# Non-invasive Transcriptomic Cell Profiling of the Human Endometrium with Generative Deep Learning

**DOI:** 10.64898/2026.05.18.26352867

**Authors:** Alvin Meltsov, Juan Manuel Falcon-Perez, Roberto Matorras, Apostol Apostolov, Alberto Sola-Leyva, Masoud Zamani Esteki, Andres Salumets, Elina Aleksejeva-Zagura

## Abstract

**Background:** Delineating the cellular origins of extracellular vesicles (EVs) enables the detection of clinically relevant changes in dynamic and complex tissues, such as the endometrium, which are not characterizable through single biomarker assays. Transcriptome deconvolution into cellular composition using deep learning methods provides a means to explore this complexity. However, such computational methods have not been previously applied to EV bulk transcriptomes, and their efficacy in profiling EV population changes and concordance to tissue throughout the menstrual cycle remains unknown.

**Methods:** This observational cross-sectional study utilized a deconvolutional generative deep learning algorithm, BulkTrajBlend, trained on a comprehensive human endometrial single-cell RNA sequencing (scRNA-seq) atlas. The model was applied to deconvolve paired bulk transcriptomes from endometrial tissue and uterine fluid EVs (UF-EVs) across the proliferative (P, n=4), early-secretory (ES, n=5), mid-secretory (MS, n=5), and late-secretory (LS, n=5) phases from healthy, fertile women. To validate generalizability, independent UF-EV datasets (ES, n=12; MS, n=12) obtained via different laboratory protocols were included. Deconvolved pseudo-single-cell (pSC) profiles from UF-EV data were subsequently integrated with Visium spatial transcriptomics slides of human endometrium (P, n=2; MS, n=4; ES, n=2).

**Results:** We developed a foundation model-based approach utilizing self-supervised learning to determine the cellular origin of EVs from their transcriptomic profiles. By mapping the generated pSC profiles to spatial transcriptomic data, we evaluated spatial origins of EVs. The statistical analysis demonstrated that UF-EV transcriptome deconvolution reflects the dynamic changes in the cellular composition of endometrial tissue across the menstrual cycle phases. The ability to distinguish accurately between proliferative and decidualizing menstrual cycle phases (ROC-AUC = 0.98) using cellular profile of deconvoluted UF-Evs transcriptome enables non-invasive profiling of endometrial tissue.

**Conclusions:** Our findings indicate the feasibility of determining endometrial tissue cellular composition using UF-EV transcriptomics. This methodology enables refined, non-invasive endometrial testing, avoiding invasive biopsy procedures. Based on deconvolution results, we are able to correlate UF-EV content to tissue, and distinguish between menstrual cycle phases. These results build toward a multifactorial screening method for abnormalities within the endometrium.

## Introduction

The endometrium, inner lining of the uterus, is critical for successful pregnancy. Ovarian hormones regulate complex changes in the endometrium at cellular and molecular levels, starting with estradiol-guided proliferation of cells in the beginning of the menstrual cycle and transitioning to progesterone-driven differentiation. This process establishes a receptive environment for embryo implantation known as the “window of implantation” (WOI), during the mid-secretory phase of the menstrual cycle [1, 2]. Despite extensive research into these physiological changes, the etiology of infertility cases remains unknown. This poses a significant challenge in reproductive medicine, affecting approximately one-quarter of couples undergoing assisted reproductive technologies [3].Furthermore, even following the transfer of a euploid embryo into a morphologically normal endometrium, implantation rates remain below 70% [4].

To address these clinical gaps, extracellular vesicles (EVs) have emerged as a significant area of interest. EVs are present in uterine fluid (UF), which can be collected alongside endometrial tissue to investigate local EV-tissue interactions or analyzed independently as a source of biomarkers for infertility [5, 6]. Moreover, the early stages of embryonic attachment and implantation may be mediated through EV-based intercellular communication [7, 8]. Consequently, EVs hold considerable promises for addressing various unmet clinical needs in the field of reproductive science [9]. Progress in separating and analyzing extracellular EVs has not fully elucidated EV subpopulations and their changing contents [10, 11]. We have previously reported that UF-derived small EVs (UF-EV) transcriptome can be used to assess the endometrial receptivity using a biomarker panel that was developed based on endometrial tissue biopsies [12, 13]. Furthermore, multiomic analysis of UF-EV cargo in parallel with paired endometrial tissue samples revealed dynamic changes throughout the menstrual cycle correlating with alterations in endometrial tissue [14].

The presence of EVs in UF and their similarity to tissue on a molecular level has been established. Nevertheless, the origins of these EVs remain unclear. Previous studies have evaluated the UF-EV profiles based on biomarker evidence [15, 16]. In a recent study, evidence was found that UF-EVs originate from epithelial and immune cells by analyzing the partial protein surfaceome of UF-EVs and the EVs derived from endometrial epithelial organoids. However, the presence of EVs originating from mesenchymal or fibroblastic cells could not be excluded due to a lack of relevant markers in the profiling approach used [14]. Therefore, while biomarker based approaches offer high confidence in cellular origin, then there is risk of false-negatives due to missing gene expression patterns [16]. Moreover, the cellular composition of the endometrium is highly complex with cell types undergoing differentiation into different subtypes due to hormonal changes [17]. This dynamic nature of endometrial tissue prompts for a novel deconvolutional approach. Deconvolving the mixed transcriptomic origin of different EVs in the UF using high-resolution reference datasets of the entire endometrium would therefore allow for the evaluation of the menstrual cycle phase and immune profile on a tissue composition level.

Deconvoluting liquid biopsies using tissue single-cell transcriptomic references faces two challenges: the accuracy with which EVs recapitulate the transcriptomic profile of the tissue of origin, and the specificity of the sequenced profile to that tissue. Larsen et al. [18] demonstrated that linear models remain sensitive to technical differences in signature matrices. Furthermore, linear deconvolutional models, such as DWLS and MuSiC, often lack generalizability in high-resolution single-cell transcriptomics, leading to inflated proportions of rare cell types or failing to deconvolute small populations [19, 20, 21]. In contrast, deep learning models like TAPE and Scaden employ autoencoder architectures to capture complex latent relationships, offering greater robustness to stochastic variation and gene expression dropout [21, 22, 23]. Similar to TAPE, BulkTrajBlend utilizes a generative computational pipeline where reference cell populations are gated to represent the bulk sample [24]. This facilitates the application of the spatial deconvolution tools cell2location and Tangram to correlate UF-EV abundance with spatial transcriptomics, which, similarly to the approach described by Liao et al. [25], allows us to validate the learned cell representation of the BulkTrajBlend model with tissue morphology [26, 27, 28].

In this study, we trained the BulkTrajBlend algorithm on a large single-cell atlas of the human endometrium and applied it to paired UF-EV and endometrial tissue bulk transcriptomes [17, 24]. We verified the applicability of our algorithm by analyzing UF-EV transcriptomes obtained independently using different methodologies [14, 29]. Furthermore, we integrated the generated scRNA-seq data from UF-EV bulk transcriptomes and mapped them onto spatial transcriptomic endometrial tissue slides to map the subpopulations of cells secreting EVs [25, 27, 28, 30]. Utilizing a deep learning deconvolutional model on paired UF-EV and tissue biopsy samples we demonstrate that EVs mirror the intricate cellular compositional changes of a complex tissue.

## Methods

### Samples

Paired endometrial tissue and UF-EV transcriptomes were acquired by our group in a previous study [14]. Samples were collected from 19 fertile healthy women across four menstrual cycle phases: proliferative (P, n=4), early-secretory (ES, n=5), mid-secretory (MS, n=5), and late-secretory (LS, n=5). UF samples were obtained using a sterile intrauterine insemination catheter (Cooper Surgical, Trumbull, CT, USA) inserted transcervical, ensuring minimal contact with the uterine fundus. Subsequently, 0.5 ml of phosphate-buffered saline (PBS, pH 7.4, Thermo Fisher Scientific, USA) was injected into the uterine cavity and further aspirated to collect the UF. The recovered volume ranged from 0.1 to 0.5 ml. Catheter and UF were visually examined to verify that no tissue was collected. UF was centrifuged for 5 min at 500g at 4 °C with fixed rotor to remove mucus/debris; UF supernatant was kept at −80 °C. UF supernatants were thawed on room temperature once, supplemented with PBS up until 0.5 ml and EVs were isolated using miniPURE-EV size-exclusion chromatography (SEC) columns under gravity according to manufacturer’s instructions (HansaBioMed Life Sciences, Tallinn, Estonia). The fractions containing particles were identified with nanoparticle tracking analysis (NTA) using sensitivity set at 85 and shutter speed at 100 and following quality control steps suggested by the manufacturer (ZetaView PMX-120 instrument, Particle Metrix, Inning am Ammersee, Germany). Fractions with particles (9–15) were pooled and directly processed. EVs presence was verified with western blot against ALIX and TSG101; multiplex bead-based EV flow cytometry against CD9, CD63 and CD81 and transmission electron microscopy (TEM). This confirmed the presence of EVs according to the MISEV2023 guidelines [31]. Detailed information on characterization methodology and data can be found in Apostolov et al. 2025 [14]. For EV-RNA isolation and western blot, EVs were precipitated from the SEC-obtained EV suspension with glycosaminoglycans-interacting reagent evGAG (HansaBioMed Life Sciences, Tallinn, Estonia), using reagent-to-sample volume ratio of 1:1, according to the manufacturer’s instructions. Two additional unpaired UF-EV transcriptomes were obtained using the same isolation protocol from two healthy donors, one from MS and another from LS cycle phase. Detailed description of RNA extraction, library preparation, and sequencing is reported in Apostolov et al. 2025 [14]. Of note, transcriptome libraries were prepared with TruSeq exome RNA library preparation kit (Illumina, San Diego, CA, USA).

UF-EV transcriptomes from Giacomini et al., 2021 [29], encompassing ES (n=12) and MS (n=12) samples from healthy fertile women were obtained as follows. UF was collected by lavaging the endometrial cavity with 2.5 ml sterile saline using a balloon hysterosonography catheter to prevent vaginal contamination and processed immediately to avoid freeze–thaw cycles. Recovered volume varied from 0.8 to 1.8 ml. UF was centrifuged at 1200g for 10 min at 4 °C to separate mucus. The supernatant was kept at 4 °C, while mucus was resuspended in PBS (pH 7.6, 1:1; Euroclone, Italy), vortexed for 3–5 min, and centrifuged at 800g for 5 min. The resulting supernatant was combined with the liquid fraction and sequentially centrifuged (300 × g, 10 min; 2000g, 20 min; 110 000g for 2 h at 4 °C, TLA-55 rotor, Optima TLX; Beckman Coulter). Pellets were resuspended in PBS, ultracentrifuged again at 110 000g for 90 min, and finally suspended in 110 µl PBS filtered through a 0.1 µm membrane and stored in sterilized 1.5-ml polypropylene tubes (Beckman Coulter Inc., Brea, CA, USA, item no. 357448). UF-EVs were characterized with NTA; western blot against CD9, CD63, ALIX, TSG101, calnexin, TOM20 and TIM44 and TEM. Together, these analyses confirmed the presence and purity of EVs consistent with MISEV2023 recommendations [31]. Detailed description of RNA extraction, library preparation, and sequencing is reported in Giacomini et al. 2021 [29]. Of note, transcriptome libraries were prepared using SMART-Seq® v4 Ultra® Low Input RNA Kit (Takara Bio Inc., Otsu, Shiga, Japan).

Proliferative and early-secretory endometrial tissue spatial transcriptomics Visium slides (P, n=2 and ES, n=2) were acquired from Garcia-Alonso et al. 2021 [27] (ENA repository E-MTAB-9260) and the MS phase (n=4) from Zhang et al. 2025b [30] (GEO repository GSE287278). As a reference for deconvolution, human endometrial cell atlas (HECA) was used by including cells from healthy donors and excluding cells from endometriosis patients and patients taking exogenous hormones, leaving 157,083 cells [17]. Due to the low number of representative cells (n<100) for ‘Glandular_secretory_FGF7’ cell state, these were combined with the ‘Glandular_secretory’ cell type.

### Quality control of transcriptomic data

Exclusion criteria were based on sample clustering characteristics and read count metrics. Three paired samples (1 UF sample from P, 1 from MS and 1 from LS phase) were removed as they were outliers per hierarchical clustering of transcript per millions (TPM) counts (Fig. S1a). For those outliers, we also detected a low amount of protein coding biotype reads from featureCounts and a high amount of reads mapping to the intronic region according to QualiMap alignments (Fig. S1b,c). As the ratio of reads marked as intronic correlated with the overall clustering of the transcriptome, we excluded three samples with more than 15% of intronic reads (Fig. S1b). One paired sample was removed due to possible switch of the UF-EV and tissue origin sample based on uniform manifold approximation and projection (UMAP) dimensionality reduction of the TPM normalized counts (Fig S1d). After quality control 15 endometrial tissue and UF-EV sample pairs and two individual UF-EV samples remained. We introduced technical replicates to detect the batch effect between sequencing libraries. All three technical replicates included in each sequencing batch demonstrated a very strong correlation between replicates (Fig. S1e, Spearman’s ρ > 0.984). From Giacomini et al. 2021 [29] all samples were included, as these samples were already considered good quality samples.

### Pre-processing of transcriptomic data

Demultiplexing was done with Illumina BCL convert v3.10.12 and preprocessed with the nf-core/rnaseq pipeline (v3.14) [32]. The reads were trimmed with Trim Galore! (v0.6.10) to remove adapter sequences and low-quality bases. Reads were aligned to the GRCh37.p13 reference genome to maintain compatibility with the Giacomini et al. 2021 preprocessed data, using the STAR aligner (v2.7.10a) [33]. Gene expression levels were quantified with the RSEM algorithm (v1.3.3) [34]. Genomic feature quantification was done with QualiMap and featureCounts (v2.0.3) [35]. Both TPM and raw reads were used in downstream analysis. Preprocessing, statistical testing and visualisation was performed with Python (v3.9 for preprocessing and v3.11 for analysis and visualisation) programming language. The HECA dataset was normalized using the preprocess function from the OmicVerse (v1.6.7) package. This process applied Pearson residual normalization, selection of the top 3,000 highly variable genes, and log-transformation of the data. Pearson residuals were chosen due to their strong performance in a recent benchmarking study by Ahlmann-Eltze & Huber, 2023 [36]. Log-transformed data underwent downstream analysis using the scanpy (v1.11.1) suite of analytical tools, including scaling, dimensionality reduction via principal component analysis (PCA) and UMAP, and the construction of a neighborhood graph of observations [37].

### Deconvolutional model parameters

The BulkTrajBlend ensemble model, consisting of the deconvolutional AE based on TAPE, a generative VAE component and a graph neural network, was configured using several key parameters for optimal performance [21, 24]. We trained the model using the HECA endometrial cell atlas [17]. To focus on the most informative features, the top 250 highly variable genes of the single cell dataset were selected. For computational efficiency, the maximum number of single cells used for training was set to one-eighth (n=19,635) of the total available single-cell profiles (n=157,083). Although initial loss plot analysis suggested convergence at around 500 epochs, the training was extended to 1,000 epochs to further decrease the reconstruction loss (Fig. S2a,b). The StandardScaler function was chosen for data scaling to harmonise feature distributions. The model was run in “high-resolution” mode to perform inference per sample. The ratio_num parameter, influencing synthetic bulk sample generation, was set to one.

We generated pseudo-single-cell (pSC) data from the bulk transcriptomic samples using the trained model. pSC data was generated using the same parameters as in the training. The pSC data was then preprocessed the same way as the scRNA-seq HECA data through normalisation, scaling, dimensionality reduction, calculating a neighborhood graph of observations and finally visualized.

### Batch correction of combined UF-EV dataset

Giacomini et al., 2021 [29] and Apostolov et al., 2025 [14] UF-EV datasets were combined and batch correction was applied with the sva package ComBat-seq function in R v3.50.0 [38]. To investigate the effect of batch correction on the deconvolution results, cell type proportions were inferred using a deep learning model on the combined dataset. This was performed both before and after applying ComBat-seq to compare the effect of batch correction with PCA dimensionality reduction.

### Cycle phase prediction based on deconvoluted cell ratios

Combined Giacomini et al., 2021 [29] and Apostolov et al., 2025 [14] UF-EV datasets were used for predicting endometrial cycle phases from cell type deconvolution fractions. The machine learning pipeline was implemented in Python using scikit-learn, evaluating a baseline Logistic Regression and a Random Forest classifier. To investigate model performance across all cycle phases we evaluated precision, recall, and F1-score using 5-fold stratified cross-validation to assess per-class performance. To focus on predicting the clinically relevant endometrial profile shift from proliferative to decidualizing, we excluded post-receptive samples and merged the P and ES classes, which increased class support and allowed for binary classification. Model performance was assessed using out-of-fold pooled predictions to calculate ROC-AUC, PR-AUC, balanced accuracy, and F1 metrics. Finally, the biological relevance of the input features was evaluated by extracting Gini impurity-based feature importances from the Random Forest model, with statistical significance established using a 100-iteration Monte Carlo label permutation test to calculate empirical p-values and a 95th percentile null distribution threshold.

### Spatial transcriptomic deconvolution and projection

Preprocessing for the spatial analysis was performed using the OmicVerse ov.pp.preprocess() function with the following parameters: mode=“pearson|pearson”, n_HVGs=3,000, log=True, and scale=True. Simulated pSC RNA-seq data from UF-EVs and endometrial tissue bulk transcriptomes, or direct scRNA-seq data from HECA were projected onto Spatial Transcriptomic Visium slides by using Tangram (v1.0.4) and cell2location (v0.1.4) with default settings [26, 28]. Projected cell abundances per spot were then inspected visually and compared to the morphology of the corresponding histological image. Overview of the workflow is visualised in Fig. 5a.

### Spatial clustering statistical testing

The accuracy of the resulting spatial cell type mapping was assessed by comparison against reference scRNA-seq HECA data using multiple metrics: mean absolute difference, calculated as the mean of absolute differences in cell type abundance between projected and reference data, centroid distance, representing the distance between the centroids of the projected and reference cell type spatial distributions; and the Jaccard index (Fig. S3). For Jaccard index, spots were assigned to the cell type exhibiting the maximum projected abundance, followed by calculation of the intersection over union with the reference cell type assignments. Pearson correlation was calculated to measure similarity of cell type abundances of pSC datasets to reference (Fig. S4). Additionally, spatial clustering was performed independently on each projected slide to evaluate consistency between. The spatial autocorrelation of the resulting spatial cell type abundance patterns was assessed using Moran’s I test. Significant Moran’s I values (p<0.05) were filtered based on a permutation test with 1,000 permutations, and FDR adjustment was applied to account for multiple hypothesis testing.

### Statistical analyses

Statistical analyses were conducted in Python 3.12 using the SciPy (v1.15.2) and NumPy (v2.2.4) packages. Welch’s t-tests, assuming unequal variances, were performed for pairwise comparisons using the ttest_ind function with equal_var=False. Tukey’s honestly significant difference post-hoc tests and one-way ANOVA were carried out using the pairwise_tukeyhsd and f_oneway functions, respectively, all with default parameters. For all tests, p-value < 0.05 was considered significant.

## Results

### Compositional changes in paired endometrial tissue and UF-EV samples across menstrual cycle

We have previously obtained paired endometrial tissue and UF-EV bulk transcriptomes from 19 fertile women across four phases of the menstrual cycle (Fig. 1a,b) [14]. The transcriptomes showed similar complexity based on RNA subtype proportions (Fig. 1c). As most samples had intronic content under 15% (n=40), we excluded those with the content exceeding that threshold due to likely poor quality (n=3) (Fig. S1b) (Table S1). To train the BulkTrajBlend deconvolution algorithm, we used an integrated single-cell reference atlas of the human endometrium, HECA, comprising 157,083 cells from healthy fertile women across all menstrual cycle phases (Fig. 1a). Although our endometrial tissue samples represent the functional layer of endometrium, we trained the model also with cell types from basal endometrium and surrounding myometrium as UF-EVs could originate from these layers due to transcytosis (Fig. 1a). Definitions of cell states and types are provided in Figure 1 caption.

**Figure 1.**
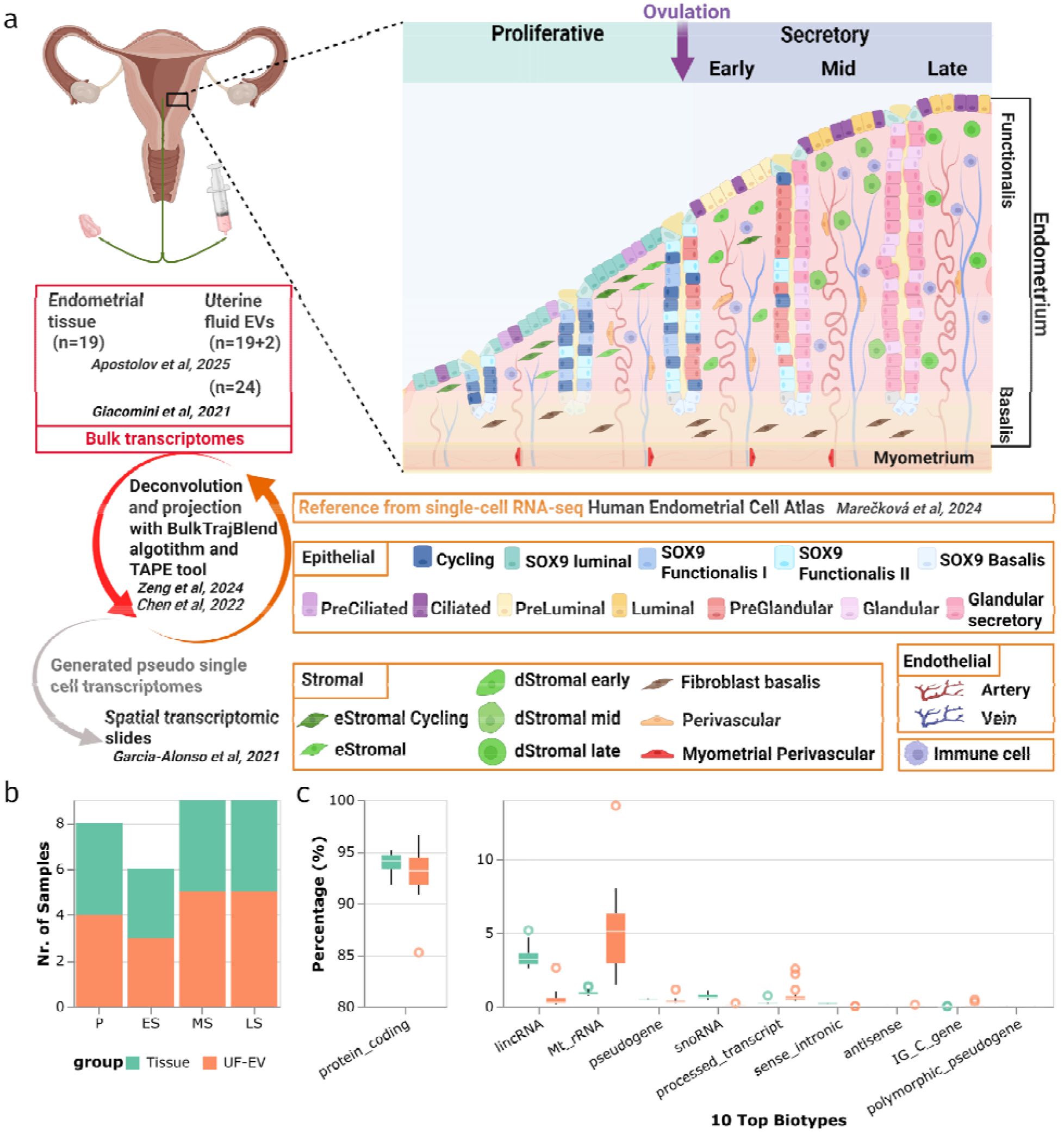
Overview of the study and datasets. **(a)** Schematic overview of the study and datasets. **(b)** Barplot showing the sample numbers of paired endometrial tissue and UF-EVs for each menstrual cycle phase. **(c)** Boxplot showing the distribution of top biotype counts in paired endometrial tissue and UF-EV transcriptomes. Abbreviations - P: Proliferative; ES: Early-secretory; MS: Mid-secretory; LS: Late-secretory; UF-EV: Uterine Fluid Extracellular Vesicles. Definitions of cell states and types: **Cycling** - epithelial cell state with high expression of cell cycle genes across glandular and luminal compartments, **SOX9 luminal** - luminal epithelial subset expressing progenitor markers, **SOX9 Functionalis I** - early differentiation stage of glandular epithelium, **SOX9 Functionalis II** - early differentiation stage of glandular epithelium, **SOX9 basalis** - progenitors responsible for regenerating the functionalis layer after menstruation, **PreCiliated** - transitional epithelial cells preparing for ciliogenesis, **Ciliated** - luminal epithelial cells that express motile cilia components, **PreLuminal** - early secretory luminal epithelial state responsive to progesterone, **Luminal** - differentiated secretory luminal epithelial cells (secrete adhesion molecules, modify glycocalyx), **PreGlandular** - early secretory glandular epithelial state responsive to progesterone, **Glandular** - differentiated mid-secretory glandular epithelium (secrete glycodelin, lipids, and other factors), **Glandular secretory** - differentiated late-secretory glandular epithelium (reparative, wound healing gene expression), **eStromal Cycling** - fibroblastic cell state with high expression of cell cycle genes, **eStromal** - proliferative phase fibroblasts, **dStromal early** - differentiated fibroblasts (decidualized), responsive to progesterone, **dStromal mid** - differentiated fibroblasts (decidualized), supports glandular epithelial maturation, **dStromal late** - differentiated fibroblasts (decidualized), premenstrual state, non-responsive to progesterone, **Fibroblast basalis** - differentiated fibroblast in basalis layer, maintains the stem/progenitor niche, **Perivascular** (ePV 1a, 1b, 2) - hormone-responsive fibroblast-like cells supporting endometrial regeneration and immune crosstalk, **Myometrial Perivascular** (mPV) - differentiated cell type supporting myometrial blood vessels.

First, we estimated the cellular composition of bulk samples and investigated the changes in cellular proportions across the menstrual cycle of the paired endometrial tissue, UF-EVs and the reference scRNA-seq dataset (Fig. 2a). For the paired samples the cellular proportions are estimated from the transcript reads, whereas for scRNA-seq the scaled cell counts per cell type in each cycle phase were used for comparison. Similar trends were observed for major cell types/states in tissue, UF-EV and reference scRNA-seq datasets. Specifically in P, the dominant cell types were SOX9 Functionalis II epithelial cells and eStromal cells; in ES, preGlandular cells; and in MS, Glandular and dStromal mid cells. However, preLuminal and Luminal epithelial cells were underrepresented in deconvoluted samples, with maximum 6% in scRNA-seq ES phase samples, whereas the maximum value over all deconvoluted samples was at 2% (Fig. 2a, Table S2). Contrary, cycling epithelial cells had higher proportions than reference, with maximum values of 11% in contrast to the maximum of 2% in reference data (Fig. 2a, Table S2). Additionally, proportional changes of transitional states of differentiated stromal cells (dStromal early/late/mid) were only partially recapitulated in deconvoluted samples (Fig. 2a). In the reference scRNA-seq dataset, the epithelial cell lineage contribution to whole tissue increased from P to LS (20% to 60%), with a concurrent decrease in the stromal cell lineage (60% to 30%), whereas uniform contributions were seen for deconvoluted samples. Remarkably, UF-EVs have a higher proportion of myeloid cells and smaller proportion of epithelial cells than tissue samples.

**Figure 2.**
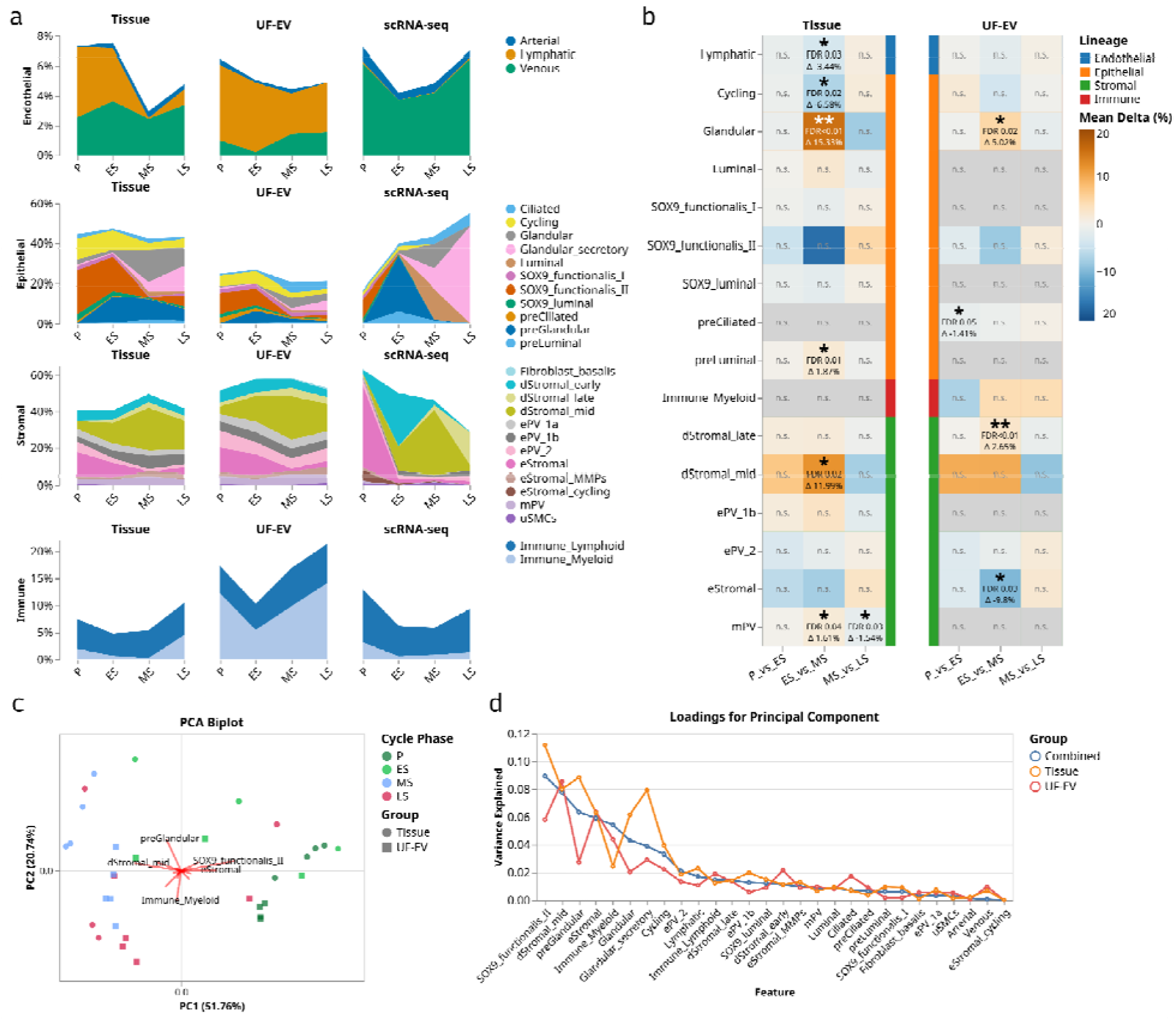
Cell proportion analysis of deconvoluted paired endometrial tissue and UF-EV transcriptomes across menstrual cycle. **(a)** Changes in cellular proportions across the menstrual cycle in endometrial tissue and UF-EVs. Abundances for each cycle phase were averaged and scaled. For visual clarity, cell types/states are divided into four categories: epithelial, stromal, endothelial and immune lineages. **(b)** Statistical analysis of cell type/state proportional changes across menstrual cycle. Cell types that showed significant changes with F-test (p < 0.05) in either endometrial tissue or UF-EV samples are marked with mean difference scale (blue/yellow), non-significant are grey. Tukey’s pairwise test results (FDR < 0.05) between consecutive menstrual cycle phases are marked with asterisks. **(c)** Biplot of PCA and largest loadings. **(d)** Factor analysis using PCA loadings of all the combinations of the datasets, PCA on endometrial tissue (orange), of combined tissue and UF-EV (blue) and of UF-EV (red). Abbreviations - P: Proliferative; ES: Early-secretory; MS: Mid-secretory; LS: Late-secretory; UF-EV: Uterine Fluid Extracellular Vesicles; scRNA-seq: Single-cell RNA-seq; PCA: Principal Component Analysis; PC1: Principal Component 1; PC2: Principal Component 2; FDR: False Discovery Rate.

Next, we tested if estimated cell abundance changes are significant across the menstrual cycle. Statistical testing was conducted both on consecutive menstrual cycle phase pairs using Tukey’s test and across the entire menstrual cycle using a F-test, p-values lower than 0.05 were considered significant. Overall, 16 cell types were considered to have significant changes throughout the menstrual cycle in both groups according to the F-test, with 8 cell types shared between UF-EV and tissue groups (Fig. 2b). In these cell types, the direction of change during the same phase was consistent between two groups. Significant increase in some differentiated cell types (dStromal mid and late, glandular epithelial cells) and a decrease in proliferating cells (eStromal and Cycling epithelial cells) were observed in both deconvoluted tissue and UF-EV samples across the menstrual cycle and for some cell types additionally between consecutive phases (Fig. 2b). Notably, the only cell type to have significant change between the same cycle phases (ES to MS) in both tissue and UF-EV was the epithelial glandular cell type, increasing from ES to MS phase by 15% in tissue and 5% in UF-EV (Fig. 2b).

Finally, we performed PCA on the deconvoluted cell proportions of tissue and UF-EV samples to explore whether the samples would cluster according to menstrual cycle phase (Fig. 2c,d). We observed a separation of samples into clusters from P and ES versus MS and LS phases, except for one tissue and UF-EV sample pair from LS, which clustered with P/ES samples. Clustering reflects the increasing effect of progesterone on cellular differentiation starting during the ES phase: P/ES samples clustering is predominantly explained by eStromal, SOX9 functionalis II cells and preGlandular cells; MS samples by dStromal mid cells and LS samples by Immune Myeloid cells (Fig. 2c,d). Variance in UF-EV samples among menstrual cycle phase groups was best explained by differentiated and proliferative stromal cells, dStromal and eStromal, respectively, and SOX9 functionalis II epithelial cells, whereas for tissue samples in addition to SOX9 functionalis II also preGlandular and Glandular secretory epithelial cells and to a lesser extent differentiated stromal cells (dStromal mid) (Fig. 2d).

### Correlation between deconvoluted endometrial tissue and UF-EV samples

To systematically assess the similarity of paired endometrial tissue and UF-EV samples, we first did hierarchical clustering on the deconvoluted cell proportions per cycle phase (Fig. 3a). UF-EV samples generally clustered apart from endometrial tissue, regardless of the phase during which the samples were collected. Notably, one tissue and UF-EV sample pair from the MS and one from LS phase clustered separately from all other samples in the same phase indicating that deconvolution captures individual differences, as these paired samples were more similar to each other than other samples in the group.

**Figure 3.**
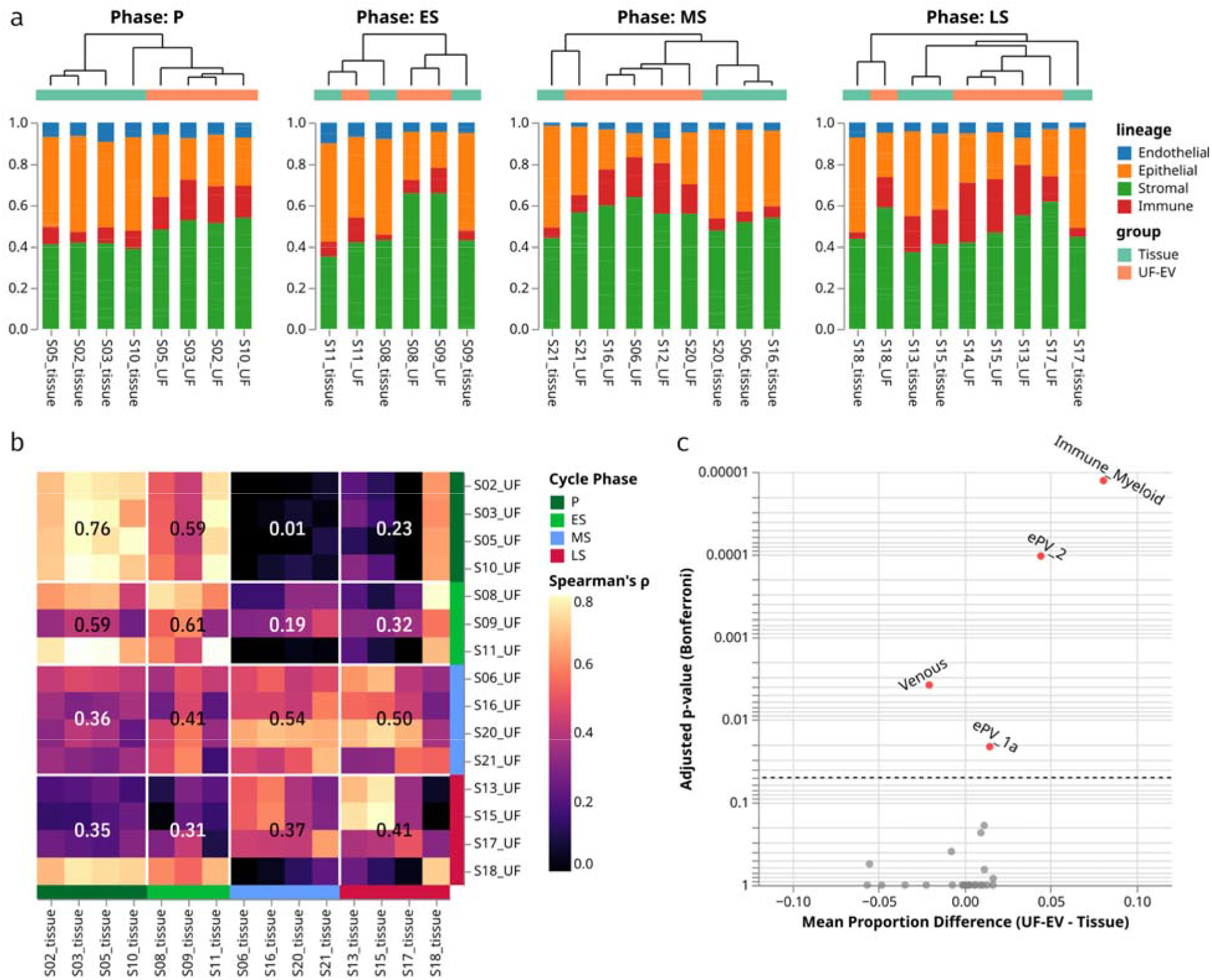
Cell type profile comparison between paired endometrial tissue and UF-EV samples. **(a)** Hierarchical clustering on deconvoluted cell proportions of paired tissue and UF-EV samples per menstrual cycle phase. For visual clarity, cell types/states are divided into four main cell type lineages: epithelial, stromal, endothelial and immune. **(b)** Heatmap visualizing Spearman’s correlation coefficient for each cell type/state between all paired tissue and UF-EV samples, average value is displayed numerically. **(c)** Welch’s t-test results with Bonferroni correction plotted against the mean difference between the abundances of the cell types. Significantly (p-adj < 0.05, grey dotted line) differing cell types are marked with red. Abbreviations - P: Proliferative; ES: Early-secretory; MS: Mid-secretory; LS: Late-secretory; UF-EV: Uterine Fluid Extracellular Vesicles; ePV: endometrial perivascular.

Next, we calculated the Spearman’s correlation coefficient for cell proportion values between all paired deconvoluted tissue and UF-EV samples as a heatmap (Fig. 3b). We also calculated the average correlation coefficient per cycle phase group. We noticed strong (ρ>0.6) average correlation between P and ES pairs of samples, and similar correlation between ES and P groups (ρ=0.56). Overall, correlation was higher within the same cycle phase group compared to outside of it (Fig 3b). The only exception was LS UF-EVs, which correlated more with MS tissue (ρ=0.50) than LS tissue (ρ=0.41). Although averaged groupwise correlation scores reflected paired group similarities, only 4 of 15 individual UF-EV pairs showed the highest correlation with their counterpart, indicating UF-EV sample shows greatest similarity to respective menstrual cycle phase tissue, but not necessarily to it’s paired tissue.

Finally, we did differential analysis on cell type abundances between combined endometrial tissue and combined UF-EV data and found that the majority (25/29) of cell types were not significantly different (p>0.05, Bonferroni correction), suggesting that endometrial tissue and UF-EV deconvoluted profiles are similar (Fig. 3c). Yet in UF-EVs, there is a significantly bigger contribution from myeloid cells (8%) and perivascular cells (ePV 2 and ePV 1a, 5% and 2% respectively), whereas venous endothelial cell proportion is on average 2.5% smaller than in tissue (Fig. 3c).

### Testing deconvolution pipeline with independent UF-EV dataset

To test the robustness and generalizability of our deconvolution pipeline, we analyzed our UF-EV data together with UF-EV bulk transcriptomes obtained by Giacomini et al. [29] consisting of 12 ES and 12 MS UF samples (Table S3). Consequently, we used only ES and MS samples from our UF-EV dataset. It is important to note that these two datasets were obtained using different laboratory methods to separate small EVs and different transcriptome library preparation reagents.

Hierarchical clustering after deconvolution shows that samples do not cluster according to technical origin, highlighting the applicability of the deconvolution pipeline on differently obtained datasets (Fig. 4a). Dimensionality reduction with PCA analysis revealed that deconvolution dramatically improved the clustering of samples according to menstrual cycle phase, and batch correction before deconvolution further increased similarities between the two independent UF-EV datasets (Fig. 4b). Spearman’s correlation analysis revealed a stronger correlation between UF-EV samples within the same cycle phase (coefficients of 0.63 and 0.57) compared to samples from different cycle groups (coefficients of 0.41 and 0.35) (Fig. 4c). Collectively, these findings confirm the robustness of our deconvolution pipeline and that representation-based learning methods could be used to capture tissue-derived profiles from UF-EV transcriptomes obtained with different EV separation methods and library preparations.

**Figure 4.**
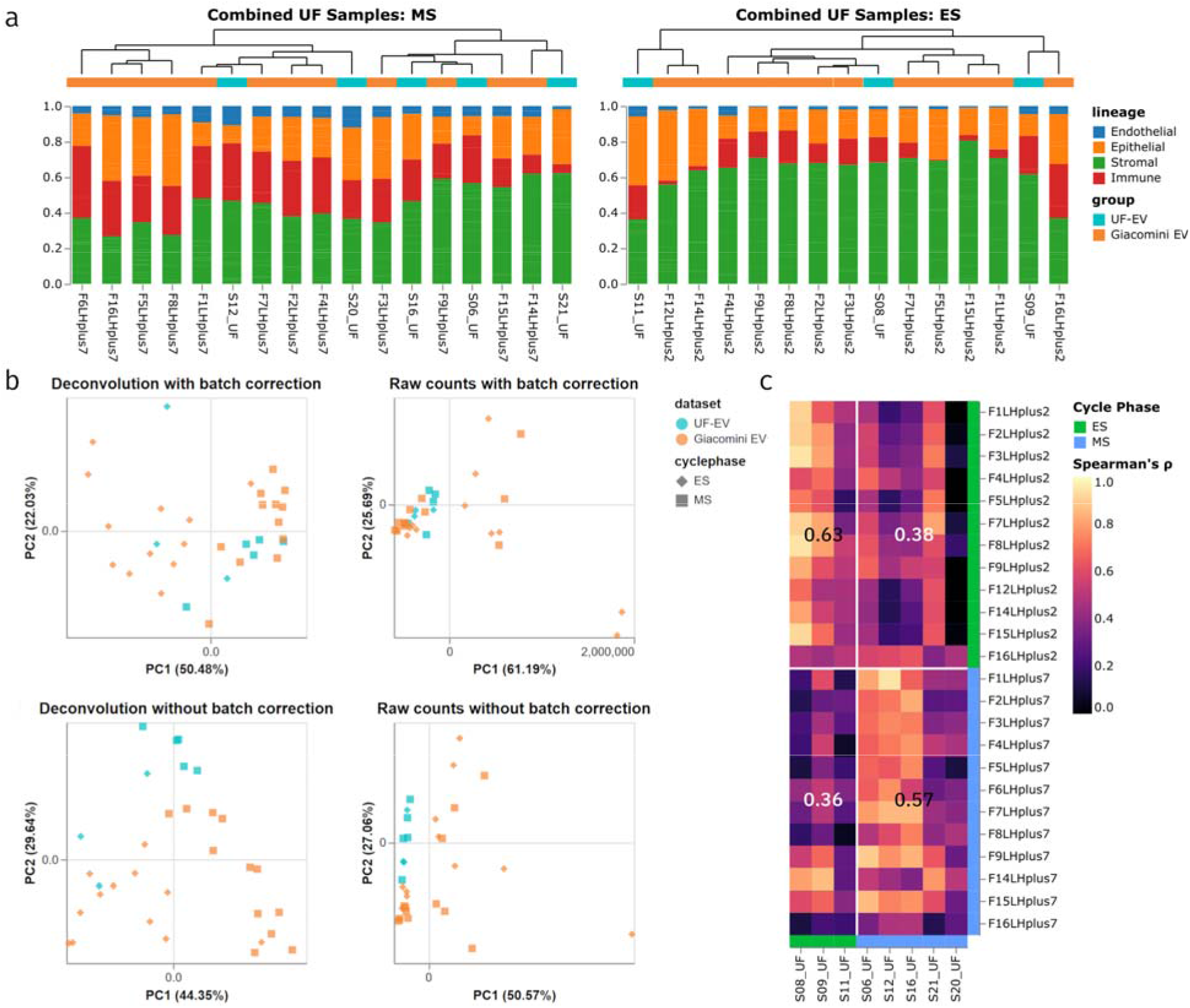
Comparison of deconvolution of two UF-EV transcriptomes obtained with different methods. **(a)** Hierarchical clustering on deconvoluted cell proportions for two UF-EV dataset obtained by Apostolov et al., 2025 and Giacomini et al., 2021. For visual clarity, cell types/states are divided into four main lineages: epithelial, stromal, endothelial and immune lineages. **(b)** PCA of two UF-EV datasets before and after deconvolution and batch correction. TPM counts were used for PCA, raw transcript counts were used for deconvolution. **(c)** Correlation heatmap between UF-EV samples obtained by Apostolov et al., 2025 and Giacomini et al., 2021. Abbreviations - ES: Early-secretory; MS: Mid-secretory; UF-EV: Uterine Fluid Extracellula Vesicles; PCA: Principal Component Analysis; PC1: Principal Component 1; PC2: Principal Component 2; TPM: transcript per million.

### Predicting endometrial cycle phase from deconvoluted UF-EV cell ratios

To see whether UF-EV-derived cellular profile could be used for endometrial receptivity assessment routinely used in assisted reproduction, we evaluated whether the combined Giacomini et al., 2021 [29] and Apostolov et al., 2025 [14] UF-EV deconvolution fractions could be used for the prediction of endometrial cycle phases.

Initial multiclass prediction using random forest and logistic regression models highlighted small sample sizes for underrepresented classes, such as P (n=4) and LS (n=5) (Table S4). Alternatively, we focused on the clinically relevant endometrial profile shift from proliferative to decidualizing stage, grouping together P and early ES as pre-receptive and keeping MS as receptive. Out-of-the fold cross validation showed robust discrimination between these two clinical groups with high AUC scores 0.98 (Fig S2a, Table S4). We extracted Gini impurity scores to determine the main contributing deconvolved cell types contributing to the prediction. This analysis showed that proliferative stromal (eStromal), fibroblast and lymphatic cell proportions are the most important discriminators between receptive and pre-receptive groups (Fig S2b).

### Spatial analysis of UF-EV and biopsy simulated datasets

To elucidate the cellular origin of UF-EVs using complementary method, we generated pSC profiles from endometrial tissue and UF-EV samples and projected them on spatial transcriptomics slides from P (n=2), ES (n=2) and MS (n=4) phases that were collected, imaged and sequenced by Garcia-Alonso et al. [27] and Zhang et al. [30] (Fig. 5a). As both simulated cell datasets from endometrial tissue and UF-EVs were deconvoluted using HECA scRNA-seq dataset as a reference, we also projected HECA dataset to the spatial transcriptomic slides and used it as a baseline reference (Fig. 5a,b).

**Figure 5.**
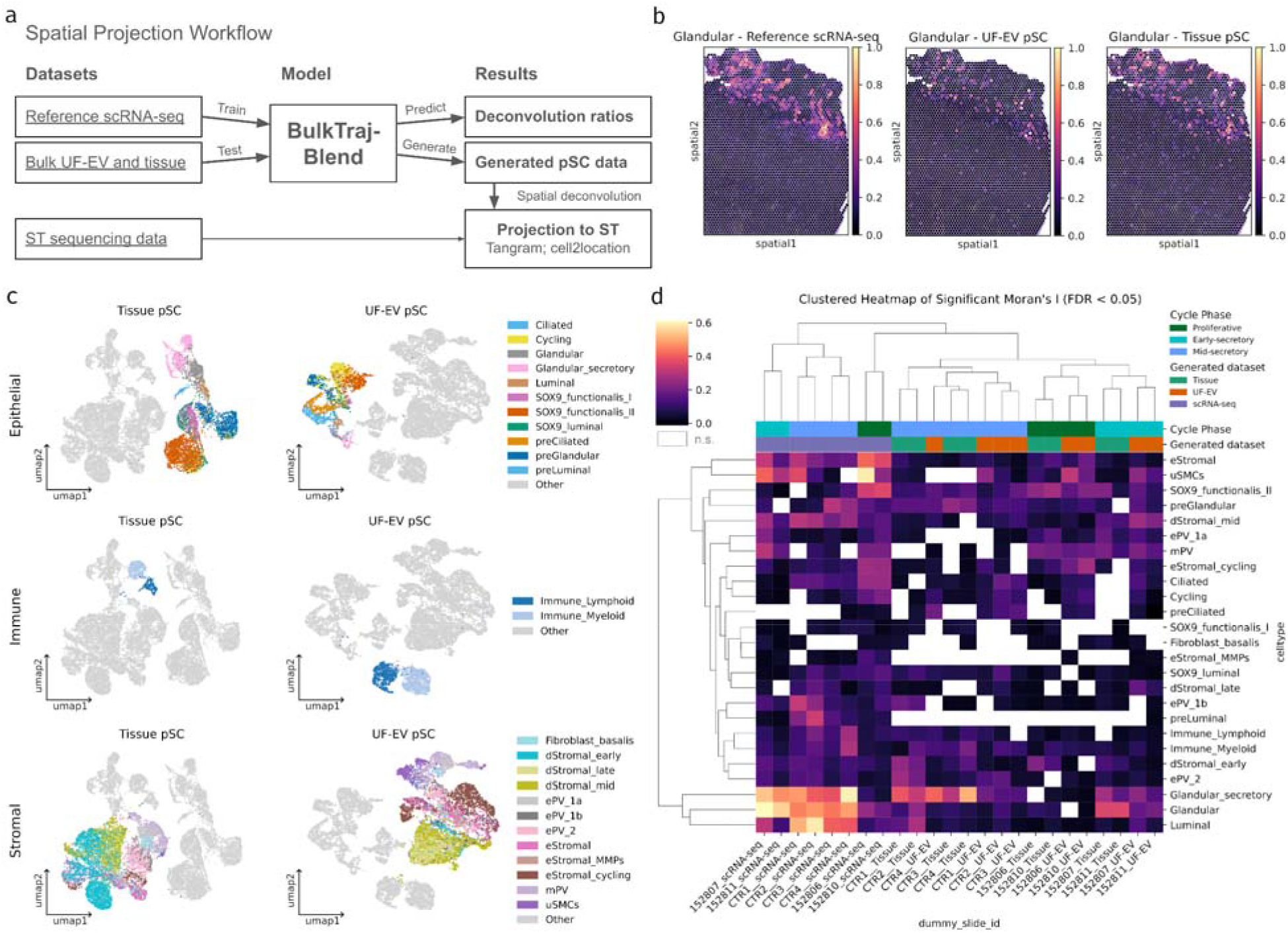
Spatial projection of simulated single cell transcriptomes based on UF-EV and endometrial tissue deconvoluted transcriptomic profiles. **(a)** Schematic representation of deconvolution, pSC data generation and spatial deconvolution analysis steps. **(b)** Projection of simulated single-cell profiles from endometrial tissue and UF-EV samples onto a ST slide (ES phase). Three glandular cell-type enrichment spatial profiles are shown using HECA scRNA-seq data, generated UF-EV pSC, and generated endometrial tissue pSC datasets. HECA scRNA-seq was projected for reference. (c) UMAP plots of generated pSC datasets (UF-EV and tissue biopsy origin). (d) Moran’s I autocorrelation scores of reference HECA scRNA-seq, generated pSC UF-EVs, and pSC of endometrial tissue projections onto ST slides (P, n=2; ES, n=2; MS, n=4). Hierarchical clustering used the farthest point algorithm with optimal leaf ordering on all Moran’s I values. Significant Moran’s I values filtered by permutation test (1,000 permutations). White boxes represent non-significance. Abbreviations – ST: spatial transcriptomics; UF-EV: Uterine Fluid Extracellular Vesicles; scRNA-seq: Single-cell RNA-seq; pSC: Pseudo-scRNA transcriptome; ST: Spatial Transcriptomics; UMAP: Uniform Manifold Approximation and Projection; FDR: False Discovery Rate.

For spatial integration, we first created a pSC dataset by averaging UF-EV and tissue bulk data across all samples. We then performed standard single-cell transcriptomic analysis on this dataset. Visual evaluation of the cell type abundance spatial profiles revealed that UF-EV and endometrial tissue projected datasets clustered within the same specific regions, as anticipated from morphology and HECA integration analysis (Fig. 5b). To measure the autocorrelation of cell type-specific abundance over all the slides and compare simulated and reference datasets, we calculated the average centroid distance, Jaccard index of the maximum cell type abundances at spot and the mean absolute difference for all the cell type abundances for P and ES phase slides (Fig. S4). Finally, Pearson correlation was calculated to measure similarity of cell type abundances of pSC datasets to the reference (Fig. S5). Based on these overall statistics, UF-EV and endometrial tissue pSC projected spatial profiles were similar across different cell lineages.

UMAP analysis revealed distinct clusters of generated cell profiles, indicating that the dataset effectively captured the heterogeneous transcriptomic landscape of reference data and projected bulk signals (Fig. 5c). Distinct clustering was observed in both tissue and UF-EV pSC datasets. However, some cell types either failed to separate from similar cell types or clustered together with other lineages. Importantly, the pSC data generated from tissue and UF-EV are not identical reflecting the slight differences in cellular proportions seen in deconvoluted samples. Therefore, the generative process maintained unique features of both transcriptomes. Next, we calculated Moran’s I, which is a measure of spatial autocorrelation and performed a permutation test with 1,000 repeats to find cell types which were significantly enriched in groups (p<0.05) (Fig. 5d). The reference HECA scRNA-seq dataset had the biggest amount of significantly scoring cell groups, whereas the generated pSC profiles from endometrial tissue and UF-EVs performed worse. Nonetheless, even using both pSC profiles, as expected from cell proportion deconvolution results, proliferating stromal (eStromal) and epithelial (Cycling) cell subtypes were significantly autocorrelated in P phase and stromal (dStromal mid) and epithelial (Glandular) cells in the ES and MS phase (Fig. 5d). Importantly, hierarchical clustering based on Moran’s I coefficient recapitulated menstrual phase transitions, supporting the biological relevance of the observed sample stratification (Fig. 5d).

## Discussion

This study demonstrates the recapitulation of endometrial tissue composition from minimally-invasive liquid biopsy samples by employing deep neural networks trained on a single-cell atlas of the human endometrium. We utilized the BulkTrajBlend deconvolutional approach which infers high-dimensional representations of cell-type-specific expression profiles from a single-cell reference dataset, estimates the proportions of cell types in bulk transcriptomic data and generates single-cell populations [24]. We trained our model using the publicly available HECA scRNA-seq atlas of endometrial tissue [17]. Applying trained models to UF-EV bulk transcriptomes allowed us to deconvolute cell type proportions and generate pSC data for projection to spatial transcriptomic slides [25, 26, 28]. Furthermore, the derived cell type fractions robustly discriminated between pre-receptive and receptive clinical profiles, achieving an out-of-fold AUC of 0.98 (Fig. S2).

We analysed paired endometrial tissue and UF-EV samples from four consecutive menstrual cycle phases. Our analysis resolved fine-grained cellular transitions, such as the emergence of preGlandular and preLuminal epithelial cells during the early secretory phase and the sequential differentiation of early dStromal into mid-secretory dStromal fibroblasts marking the progression from estrogen-driven proliferation to progesterone-dependent differentiation [27]. We tracked the significant downregulation of SOX9 functionalis progenitors as the tissue exited the proliferative phase [17]. These shifts in UF-EV cell type ratios mirrored the synchronised differentiation of epithelial and stromal compartments observed in paired tissue samples and were confirmed to be statistically significant in both temporal and spatial dimensions (Fig. 2b, 5d). Furthermore, strong correlation was measured between UF-EVs and endometrial tissue within the same cycle phase group (Fig. 3b), validating that the liquid biopsy cargo reflects the dynamic changes in the tissue. Our findings indicate that UF-EVs recapitulate the whole transcriptome profiles of the endometrium with comparable resolution to the tissue samples.

The robustness of this deconvolution pipeline was further verified using independently obtained UF-EV bulk transcriptomic data from the ES and MS phases. Comparison with our primary dataset confirmed that the accuracy of the model is not significantly affected by differences in EV separation or library preparation methods (Fig. 4c) [14, 29]. Although ultracentrifugation is expected to retain ribonucleoproteins more than size-exclusion chromatography, and these methods may enrich slightly different EV populations, our results suggest that the retrieved transcriptomic signatures are comparable [39-42]. Notably, deconvolution improved the clustering of samples according to the menstrual cycle phase compared to bulk transcriptomic signatures alone, indicating that this approach delineates the biological state despite technical bias (Fig. 4b). Moreover, spatial colocalization of the same cell types within the same morphological niches demonstrates that the representational learning ability of the generative model can also be successfully transferred to spatial omics (Fig. 5b,d, S4, S5).

Despite the similar transcriptomic signature to the reference tissue, specific cell type proportions in UF-EVs occasionally diverge from the reference tissue profiles. These discrepancies likely reflect biological differences between cellular retention and vesicle secretion or stability. For instance, myeloid lineages appear more abundantly in UF-EVs than in tissue references (Fig. 2d, 3c). This is consistent with flow cytometry data suggesting that immune compartments have higher EV secretion rates, leading to their overrepresentation in the deconvoluted cell proportions [14]. Deconvolution results further suggest that fibroblast-like stromal and endothelial cells contribute to the UF-EV pool. This indicates that despite the immune background, the structural components of the tissue are effectively captured. We also observed a “trailing off” effect where certain cell signals, such as preGlandular and SOX9 functionalis II, persist in UF-EVs during subsequent cycle phases even after disappearing from the tissue reference (Fig. 2a). In blood circulation with rapid clearance by the liver and spleen, injected EVs might be detectable only up to 30 minutes [43, 44] or up to 5.5 hours [45]. However, the uterine lumen represents a semi-closed mucosal system where the kinetics of clearance are governed by mucosal turnover and reabsorption by the uterine epithelia. Additionally, longer half-life of EV’s may be advantageous for reproduction, allowing continuous interaction of endometrium with embryo and sperm cells [7, 46-49]. This could translate into longer retention times of EVs secreted from stromal and epithelial tissues during the proliferative and early secretory phase. Nevertheless, for some cell types, such as SOX9 functionalis II and dStromal early, persistence is observable in both tissue and UF-EV proportions. Moreover, proliferative stage specific cell types, such as preGlandular, persist throughout the cycle phases, requiring EV’s to persist for several days from proliferative to late-secretory stage (Fig 2a). This indicates that instead of capturing temporal contamination, there might be misclassification of similar cell types of epithelial and stromal origin, especially for cell types that also cluster together in the pSC datasets (Fig 5c). Both deconvolution and spatial EV origin analysis results suggest that some unexpected cell types like smooth muscle cells contribute to the UF-EV pool (Fig 2a). While their absence in paired tissue likely results from biopsy sampling biases that exclude deep-tissue lineages, their presence in UF-EVs suggests that uterine fluid captures a more complete view of the endometrium.

Our study highlights several limitations and potential avenues for future research. The performance of the BulkTrajBlend deconvolution model warrants further benchmarking against established methods like CIBERSORTx, MuSiC, and DWLS [20, 50, 51], particularly for the detection of rare cell types and highly similar subpopulations [21]. Cell mixture benchmarking setup, similar to Larsen et al. [18] with urine samples and renal tissue cell lines, should be performed with endometrial tissues, and subsequently compared with in vitro results. This benchmarking method could help explain the loss of luminal cell signals in deconvolution (Fig 2a). When analysing the spatial autocorrelation of EV biogenesis markers with cell types, we noticed that overall, absolute Moran’s I values were still relatively low for reference scRNAseq spatial deconvolution values (max: 0.6; min: -0.02) (Fig 5d). For some cases with high average Moran’s I values, such as Glandular (I=0.6), the cellular enrichment profiles were spatially well defined to their respective tissue regions (Fig 5b, d). Lower average autocorrelation values might indicate poorly localised expression of the biomarker genes for given cell type. Alternatively, this might be caused by sparse spatial coverage of gene expression due to low tissue coverage of the capture area. We observed that Visium slides from Zhang et al. [30] of the MS phase had less of the capture area covered by tissue than Garcia-Alonso et al. [27], and contained visibly less endometrial basalis region. When compared to other phases, this could lead to overall low autocorrelation values in the MS phase. These shortcomings highlight a need for development for autocorrelation methods that account for this sampling heterogeneity. Therefore, whether the overall low autocorrelation values are due to expression dropout effect in the spatial transcriptome panel or due to low EV formation in those tissues, requires further evaluation and more high quality spatial transcriptomic datasets.

Another limitation of the current analytical pipeline is that the deconvolution and generated pSC profiles are limited to the cell type composition and sampling bias of the reference single cell atlas, with no information shared between the generative and deconvolution model other than cell type ratios. A significant constraint remains the dependence on the tissue-derived reference atlas. This limitation could be remedied by sampling a whole organism single cell atlas for adjacent cell populations using resources such as the Human Cell Atlas [52]. Nevertheless, while compiling a liquid-biopsy-specific reference atlas from nearby tissue sources could theoretically improve specificity and functionality, this approach risks exacerbating batch effects and introducing off-target tissue types that resemble reference populations, potentially leading to the over-estimation of major cell types [19-21, 53]. Therefore, this study relies on the assumption that UF-EVs predominantly originate from the nearby tissue, in this case the endometrium. Future research should focus on developing methods that account for unknown profiles in bulk tissue or increasing the specificity of cell type detection to mitigate false positive or inflated cell proportion estimation for rare cell types. Additionally, more research is needed investigating the clearance and uptake time of EV’s in the uterine fluid, as this could introduce a local temporal contamination of previous cell stages of the differentiating endometrium.

Looking forward, the clinical utility of this analytic approach could be significantly expanded by incorporating scRNA-seq data from pathological conditions. Studies on recurrent implantation failure and endometriosis have identified associations of specific immune and stromal cell subclasses with implantation failure and endometrial abnormalities [54, 55]. Alternatively, EV’s might serve as carriers of biomarkers for deep-tissue pathologies such as adenomyosis [56]. Integrating these pathological cell signatures into the reference model would contribute towards developing non-invasive “all-in-one” tests [57]. Such tools could generate risk scores for various endometrium-related pathologies from a single transcriptomic profile, potentially reducing the need for invasive testing in IVF and gynecological patients. While in this study we focused on correlative exploratory analysis on using UF-EV’s and deep learning methods for non-invasive endometrial profiling and achieved high prediction accuracy (AUC = 0.98) (Fig S2), then further validation of UF-EV origin and content using larger sample sets, including larger P and LS cycle reference, and novel statistical methods should be developed before translating these results into clinical medicine [58-60].

In conclusion, we propose a deconvolution pipeline trained with reference tissue scRNA-seq data that deconvolutes bulk transcriptomes. Additionally, we explore a visual representation approach of these deconvoluted profiles by projecting generated pSC datasets to publicly available spatial transcriptomes of the endometrium and correlate these results with morphological features. We provide evidence that UF-EVs reflect dominant trends in endometrial tissue cellular composition across the menstrual cycle through temporal and spatial analysis, providing high predictive potential for non-invasive endometrial testing.

## Data Availability

The datasets supporting the conclusions of this article are available in the ArrayExpress repository (www.ebi.ac.uk/arrayexpress), E-MTAB-15505 (https://www.ebi.ac.uk/biostudies/ArrayExpress/studies/E-MTAB-15505) and GitHub (www.github.com) repository, CellularGenomicMedicine/endo-ev (https://github.com/CellularGenomicMedicine/endo-ev).

## Abbreviations

EV: Extracellular Vesicles
pSC: Pseudo-scRNA transcriptome
scRNA-seq: single-cell RNA sequencing
ST: Spatial Transcriptomics
UF: Uterine Fluid
UF-EV: Uterine Fluid Extracellular Vesicles
VAE: Variational Autoencoder
dStromal: differentiated fibroblasts (decidualized)
ePV: endometrial perivascular
ES: Early-secretory
eStromal: proliferative phase fibroblasts
FDR: False Discovery Rate
LS: Late-secretory
mPV: myometrial perivascular
MS: Mid-secretory
P: Proliferative
PCA: Principal Component Analysis
TPM: transcript per million
UMAP: Uniform Manifold Approximation and Projection

## Declarations

### Ethics approval and consent to participate

The study was approved by the Research Ethics Committee of the University of Tartu, Estonia (No. 330M-8), and written informed consent was obtained from all participants.

### Consent for publication

Not Applicable

### Author’ contributions

**Alvin Meltsov:** Conceptualization, data curation, formal analysis, investigation, methodology, validation, visualization, writing – original draft. **Juan Manuel Falcón:** Writing – review and editing. **Roberto Matorras:** Writing – review and editing. **Apostol Apostolov:** Writing – review and editing. **Alberto Sola-Leyva:** Writing – review and editing. **Masoud Zamani Esteki:** Methodology, supervision, project resources, writing – review and editing. **Andres Salumets:** Conceptualization, supervision, project resources, writing – review and editing. **Elina Aleksejeva-Zagura:** Conceptualization, supervision, validation, investigation, writing – original draft.

## Acknowledgements

The authors would like to thank Prof. Han Brunner for insightful discussions and helpful suggestions.

