## Supplementary Figures for "Non-invasive Transcriptomic Cell Profiling of the Human Endometrium with Generative Deep Learning"

[a] Celvia CC, Tartu, Estonia.

[b] Department of Clinical Genetics, Maastricht University Medical Centre+, Maastricht, The Netherlands.

[c] Department of Genetics and Cell Biology, GROW School for Oncology and Developmental Biology, Maastricht University, Maastricht, The Netherlands.

[d] Exosomes Laboratory and Metabolomics Platform, Center for Cooperative Research in Biosciences (CIC bioGUNE), Basque Research and Technology Alliance (BRTA), Derio, Spain.

[e] Ikerbasque, Basque Foundation for Science, Bilbao, Spain.

[f] Centro de Investigación Biomédica en Red de Enfermedades Hepáticas y Digestivas (CIBERehd), Madrid, Spain.

[g] Human Reproduction Unit, Cruces University Hospital, Baracaldo, Vizcaya, Spain.

[h] Department of Gynecology, Basque Country University, Lejona, Vizcaya, Spain.

[j] Department of Gynecology, Basque Country University, Biobizkaia, Baracaldo Viacaya, Spain.

[k] Department of Gynecology, Basque Country University, IVI Bilbao, IVIRMA, Lejona, Spain.

[l] Division of Obstetrics and Gynecology, Department of Clinical Science, Intervention and Technology (CLINTEC), Karolinska Institute, Stockholm, Sweden.

[m] Department of Gynecology and Reproductive Medicine, Karolinska University Hospital, Stockholm, Sweden.

[n] Department of Biotechnology, Institute of Molecular and Cell Biology, University of Tartu, Tartu, Estonia.

[o] Department of Obstetrics and Gynecology, Institute of Clinical Medicine, University of Tartu, Tartu, Estonia.

* These authors contributed equally to this work

**Running title:** Deep deconvolution for UF-EV RNA-seq data

**Contacts:** Alvin Meltsov -; Juan Manuel Falcón-Perez -, Roberto Matorras -,

Apostol Apostolov -, Alberto Sola-Leyva -; Masoud Zamani Esteki -, Andres Salumets -, Elina Aleksejeva-Zagura -

**ORCID: Andres Salumets -** 0000-0002-1251-8160

### Supplementary Materials

**Supplementary Table 1. Qualimap results of endometrial tissue and uterine-fluid transcriptomes.**

**Supplementary Table 2. Predicted cell proportions of deconvoluted endometrial tissue and uterine fluid extracellular vesicles transcriptomes.**

**Supplementary Table 3. Predicted cell proportions of deconvoluted uterine fluid extracellular vesicles transcriptomes from two different studies.**

**Supplementary Table 4. Accuracy metrics of multinominal and binary prediction models for menstrual cycle phase.**

**Supplementary Figure 1. Quality control of raw transcriptomic samples**

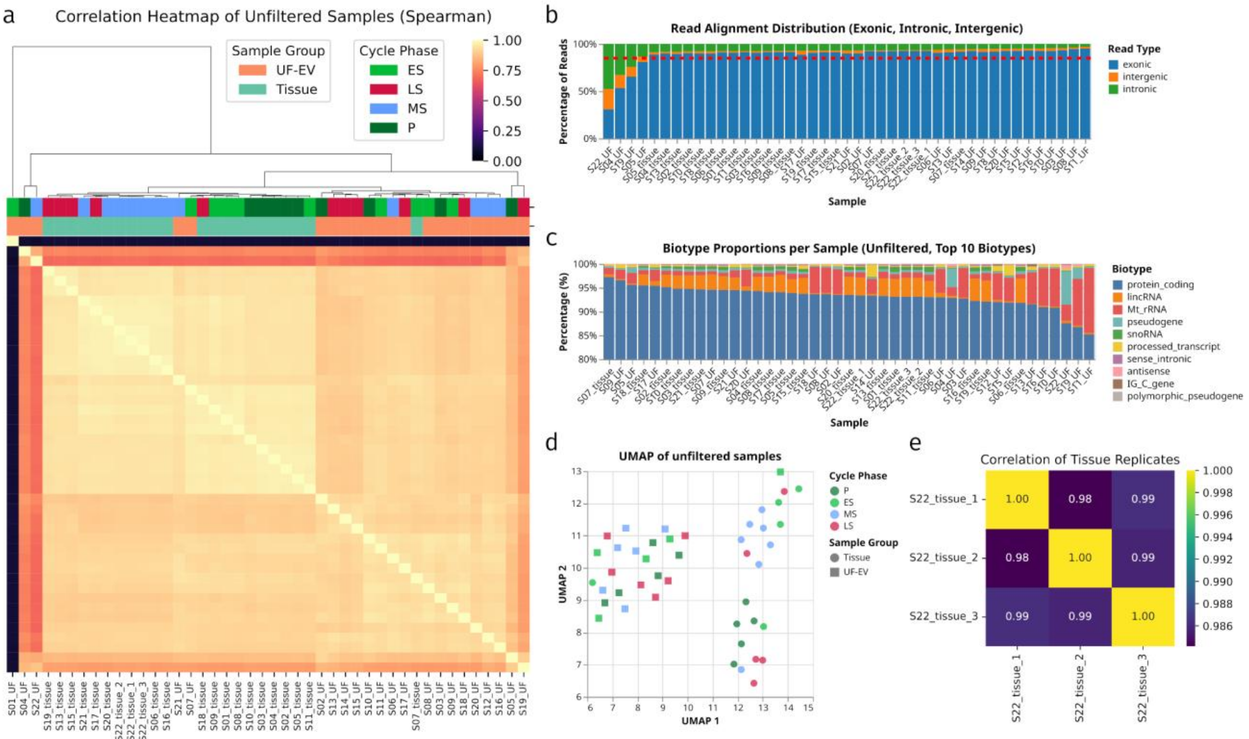


**(a)** The correlation matrix of TPM reads comparing all the samples before filtering out low quality samples. **(b)** Stacked bar plot displaying the percentage of reads aligning to exonic, intronic, and intergenic regions for each unfiltered sample. **(c)** Stacked bar plot displaying the percentage of top 10 biotype proportions for each unfiltered transcriptomic sample. **(d)** UMAP plots of samples before filtering by quality control. Menstrual cycle phase is represented with color, the type of sample with shape. **(e)** The correlation matrix of TPM reads comparing three technical replicates from different sequencing batches.

Abbreviations - TPM, Transcripts Per Million; UMAP, Uniform Manifold Approximation and Projection; UF-EV, Uterine Fluid Extracellular Vesicles; P, Proliferative; ES, Early-Secretory; MS, Mid-Secretory; LS, Late-Secretory.

**Supplementary Figure 2. Prediction metrics and feature importance values of predicting endometrial cycle phases**

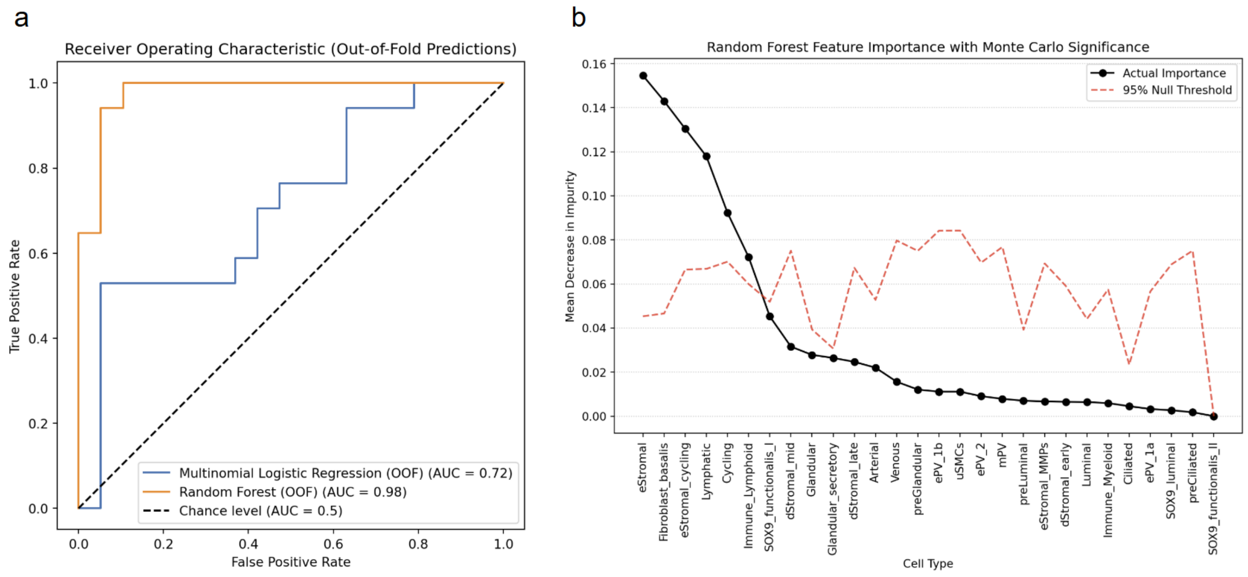


(a) Out-of-fold Receiver Operating Characteristic (ROC) curves for the binary classification distinguishing proliferative from decidualizing profiles. The Random Forest model demonstrates robust discrimination with an Area Under the Curve (AUC) of 0.98, outperforming the Multinomial Logistic Regression baseline (AUC = 0.72). (b) Scree plot detailing Random Forest feature importances quantified by the mean decrease in impurity. The solid line denotes actual importance scores for each deconvoluted cell fraction. The dashed line illustrates the 95th percentile null distribution threshold established through Monte Carlo label permutation testing. Cell types where the actual importance exceeds this empirical threshold represent statistically significant drivers of the phase classification.**Supplementary Figure 3. Preprocessing data distribution and loss plots for the deconvolution model training**

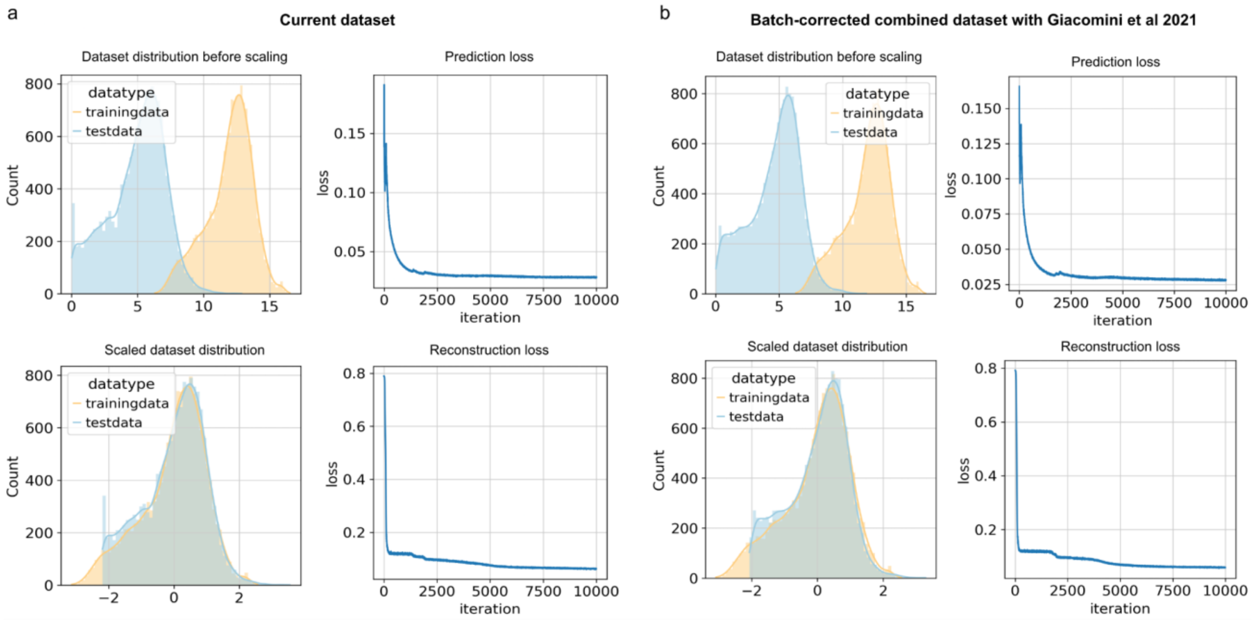


**(a)** Test data is paired endometrial tissue and UF-EV samples. The top-left panel shows the distributions of the datasets before scaling. The top-right panel displays the prediction loss over 10,000 iterations. The bottom-left panel shows the scaled distributions of training and test datasets, and the bottom-right panel illustrates the reconstruction loss over the training iterations. **(b)** Equivalent metrics for the model with batch-corrected combined test data from Giacomini et al., 2021 and Apostolov et al., 2025. The panel organization is identical to (a), showing data distributions before and after scaling and loss plots for prediction and reconstruction.

**Supplementary Figure 4. Spatial Clustering Metrics**

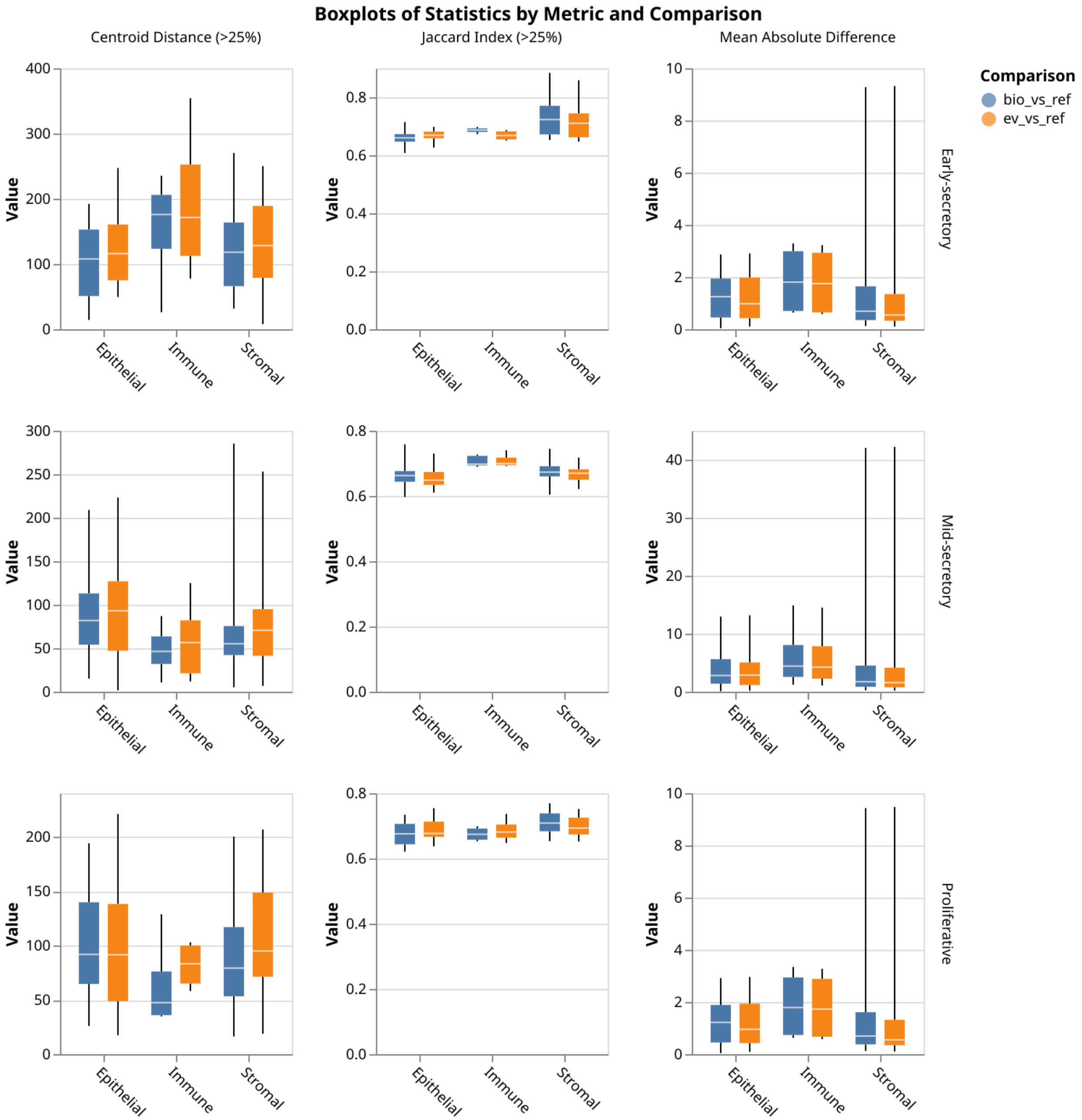


Boxplots of spatial projection statistics comparing generated pSC datasets (endometrial tissue-derived and UF-EV-derived) against the reference scRNA-seq dataset. Metrics are calculated for epithelial, immune, and stromal lineages on proliferative (bottom row), early-secretory (top row) and mid-secretory (middle row) phase slides. The metrics include Centroid Distance, assessing the distance between the spatial centroids of projected and reference cell types; Jaccard Index, measuring the overlap of spot assignments for the most abundant cell type; and Mean Absolute Difference, quantifying the average difference in cell type abundances. Metrics are calculated for all eight slides, consisting of two proliferative stage tissue samples (lower), two early-secretory stage tissue samples (top) and four mid-secretory stage tissue samples (middle).

**Supplementary Figure 5. Spatial Clustering Pearson correlation**

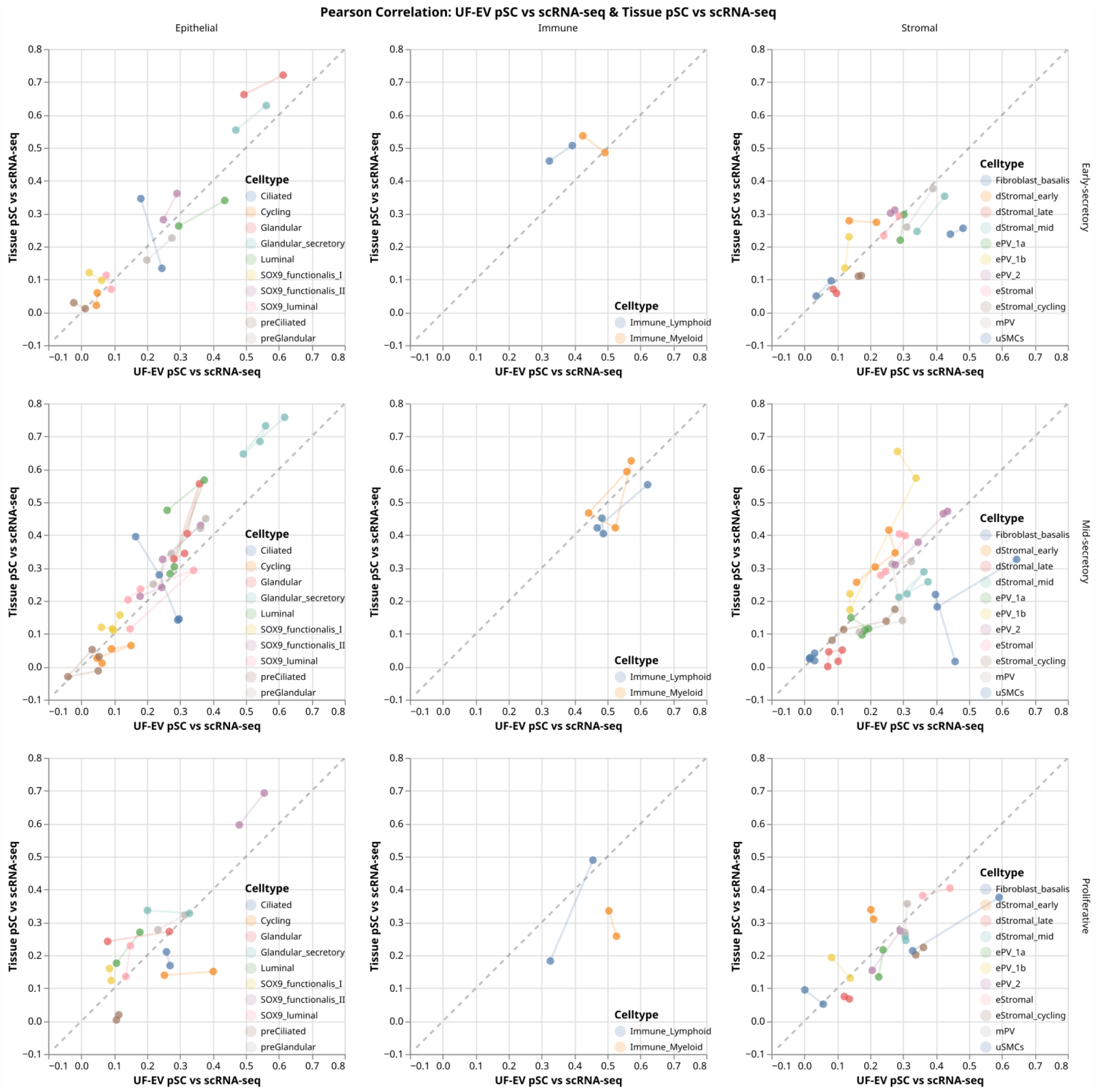


Pearson correlation of cell type abundances between pSC datasets (UF-EV pSC vs. scRNA-seq and Tissue pSC vs. scRNA-seq) for epithelial, immune and stromal lineages. The analysis is shown separately for proliferative (bottom row), early-secretory (top row) and mid-secretory (middle row) phase spatial transcriptomic slides, with the same cell types on different slides connected with a line. The dashed line represents the identity line (y=x).

Abbreviations – pSC, pseudo-single-cell; scRNA-seq, single-cell RNA sequencing; UF-EV, Uterine Fluid Extracellular Vesicles.
